# HIV as a Host Susceptibility State for Severe Drug Hypersensitivity: Disentangling Biological Susceptibility from Drug Exposure in the FAERS Database

**DOI:** 10.64898/2026.07.07.26356279

**Authors:** Eric Mukherjee, Dodi Park, Amir Asiaee, Matthew S. Krantz, Cosby A. Stone, Michelle Martin-Pozo, Elizabeth J. Phillips

## Abstract

**Background:** HIV infection has long been associated with increased incidence of severe cutaneous adverse reactions (SCAR). It remains unknown whether this increased incidence is a direct biological result of HIV infection, differences in drug exposure, or other demographic factors.

**Objective:** To evaluate the association between HIV and SCAR and determine whether this relationship persists after adjusting for demographic factors and structured drug exposure.

**Methods:** We analyzed reports from the FDA Adverse Event Reporting System (FAERS) from 2013-2023. SCAR outcomes included Stevens-Johnson syndrome/toxic epidermal necrolysis (SJS/TEN), drug reaction with eosinophilia and systemic symptoms (DRESS), acute generalized exanthematous pustulosis (AGEP), and generalized bullous fixed drug eruption (GBFDE). HIV status was determined using antiretroviral exposure, indication text, and machine-learning imputation. Logistic regression models were constructed sequentially: unadjusted, demographic-adjusted, and fully adjusted with drug principal components to account for polypharmacy. Drug-level disproportionality and HIV-drug interaction analyses were also performed.

**Results:** In unadjusted models, HIV was strongly associated with SCAR (OR ∼2.0-2.7). Adjustment for demographics attenuated this association, and further adjustment for drug exposure reduced the effect to near null for overall SCAR and DRESS. A modest residual association persisted for SJS/TEN (OR ∼1.3). Disproportionality analyses demonstrated enrichment of specific high-risk drugs in PLWH. Interaction modeling revealed drug-specific amplification of SCAR risk in HIV, notably for carbamazepine and clarithromycin, whereas other drugs showed minimal interaction.

**Conclusion:** The association between HIV and SCAR is largely explained by differences in drug exposure and demographic factors. Residual risk is drug-specific rather than uniform, supporting a model in which HIV modifies susceptibility to select drug triggers rather than acting as a global risk factor. Further prospective and retrospective studies are required to quantify associations.

**Highlights Box:** *What is already known about this topic?:* HIV infection is associated with increased risk of severe cutaneous adverse reactions, but the relative contributions of biological susceptibility and drug exposure remain unclear.

*What does this article add to our knowledge?:* This study demonstrates that much of the HIV-SCAR association is explained by drug exposure patterns, with residual risk limited to specific drugs and phenotypes.

*How does this study impact current management guidelines?:* These findings support focusing risk mitigation on specific high-risk drugs in HIV rather than assuming uniformly elevated SCAR risk across all medications.

## Introduction

Severe cutaneous adverse reactions (SCAR), including Stevens-Johnson syndrome/toxic epidermal necrolysis (SJS/TEN), drug reaction with eosinophilia and systemic symptoms (DRESS), and acute generalized exanthematous pustulosis (AGEP), are among the most severe forms of delayed drug hypersensitivity. Although individually uncommon, these syndromes are associated with substantial morbidity, mortality, and healthcare utilization worldwide.^1,2^ Despite advances in understanding the immunopathogenesis of SCAR, predicting which patients will develop severe drug hypersensitivity remains a major clinical challenge.

HIV infection has consistently been associated with an increased risk of drug reactions, with prior studies reporting several-fold higher incidence in persons living with HIV (PLWH) compared with the general population.^3^ However, the mechanisms underlying this association remain incompletely understood. HIV infection is characterized by profound immune dysregulation, chronic inflammation, altered T-cell homeostasis, and frequent viral co-infections, all of which may plausibly influence susceptibility to drug hypersensitivity. At the same time, PLWH experience markedly different medication exposures than the general population, including lifelong antiretroviral therapy, treatment of opportunistic infections, and increased polypharmacy. These differences complicate interpretation of observed associations and raise the possibility that some portion of the apparent HIV-associated risk reflects differences in medication exposure rather than intrinsic biologic susceptibility.

Distinguishing between these possibilities is clinically important. If HIV confers a generalized increase in susceptibility to severe drug hypersensitivity, heightened caution may be warranted across a broad range of medications. Conversely, if excess risk is largely attributable to specific drug exposures, targeted risk mitigation strategies may be more appropriate. More broadly, understanding how HIV modifies drug hypersensitivity may provide insight into the interplay between host immune state and drug-specific risk factors in SCAR pathogenesis. If HIV acts as a generalized amplifier of hypersensitivity, elevated risk would be expected across many medications and phenotypes, even after accounting for differences in drug exposure. In contrast, if excess risk is primarily driven by prescribing patterns and treatment complexity, adjustment for medication exposure should substantially attenuate observed associations. A third possibility is that HIV selectively modifies susceptibility to specific drug-host interactions, resulting in amplification of risk for certain medications but not others. Distinguishing among these competing explanations has proven difficult in traditional epidemiologic studies, which are often limited to individual medications, specific clinical settings, or relatively small numbers of SCAR events.

Large-scale pharmacovigilance databases provide a complementary approach for studying these questions. Although spontaneous reporting systems cannot directly estimate incidence, they capture large numbers of rare adverse drug reactions across diverse medications, populations, and clinical contexts. The FDA Adverse Event Reporting System (FAERS) contains millions of reports and is particularly well suited for evaluating SCAR, which are individually uncommon but collectively represented across a wide range of drugs and patient populations.^4,5^ Furthermore, the breadth of FAERS enables simultaneous assessment of drug-specific effects, host factors, and potential effect modification across multiple SCAR phenotypes.

In this study, we leveraged FAERS to evaluate the relationship between HIV infection and SCAR. We integrated machine learning-based HIV classification, structured modeling of medication exposure, and drug-level interaction analyses to determine whether HIV independently increases susceptibility to SCAR or whether observed associations are primarily explained by differences in medication exposure. By disentangling biological susceptibility from drug exposure, we sought to better define the role of HIV as a host susceptibility state for severe drug hypersensitivity.

## Methods

### Data Source and Study Design

All analysis was conducted in R version 4.4.2 (2024-10-31 ucrt), using RStudio 2026.05.0 Build 218. We conducted a retrospective pharmacovigilance study using a deduplicated version of the FAERS from January 2013 through December 2023.^6,7^ SCAR were defined using standardized MedDRA preferred terms and included the following phenotypes: SJS/TEN, DRESS, AGEP, and generalized bullous fixed drug eruption (GBFDE).^4^ Each outcome was analyzed separately as a binary variable at the report level. Age was imputed using hot-deck imputation based on the top 20 variables correlated with age (simputation package v0.2.9).

### Drug Exposure Definitions

Drug exposures were defined using FAERS role codes, with primary suspect (PS) drugs representing the main exposure of interest. Drug names were mapped to standardized concept identifiers, and combination products (e.g., trimethoprim-sulfamethoxazole) were fissioned.

### HIV Classification and Imputation

HIV status was determined using a structured, multi-stage approach integrating antiretroviral (ARV) exposure, indication data, and model-based imputation (**Figure S1**). Drug names were standardized and mapped to a curated reference list, and binary indicators were generated for overall ARV exposure, class-level exposure, and individual drug exposure. To derive an indication-based HIV proxy, drug and indication tables were merged at the patient-drug level, and all indications associated with ARV medications were extracted. Indications were manually annotated as HIV-positive, non-HIV indication, probable HIV, or ambiguous. These labels collapsed to the patient level using a hierarchical rule: patients were classified as HIV-positive if any indication explicitly reflected HIV care, as non-HIV if only non-HIV indications were present, and as ambiguous otherwise. Additionally, indications labeled as probable HIV were not used to assign definitive HIV status but were retained as a separate feature capturing indirect clinical evidence of HIV infection, such as treatment patterns or HIV-associated conditions. This allowed preservation of clinically meaningful uncertainty without forcing premature classification.

Reports with ambiguous indication data underwent imputation using missForest (v1.5), a nonparametric random forest-based imputation algorithm. The imputation model included age, sex, geographic region, medication burden, death, antiretroviral exposure variables at the overall, class, and individual-drug levels, an HIV-probable indication flag, and DrugPCs summarizing global medication exposure patterns. Adverse event outcome variables were excluded from the imputation model to avoid circularity. Definite HIV-positive and definite non-HIV labels were preserved, and only ambiguous labels were updated by imputation. The model was run with 100 trees and up to 5 iterations, with a fixed random seed.

### Drug-Level Disproportionality Analysis

To identify drugs disproportionately associated with SCAR in HIV-positive patients, we calculated reporting odds ratios (RORs) using primary suspect designations within the HIV-positive population. Analyses were restricted to drugs meeting minimum case thresholds ≥5 cases among HIV-positive reports). Continuity corrections were applied where necessary. False discovery rate (FDR) adjustment was used to account for multiple comparisons.

### Drug Principal Components and Logistic Regression

To account for confounding arising from complex patterns of medication use, we generated a sparse patient-by-drug exposure matrix using all reported drug exposures. Drugs reported in fewer than 100 patients were excluded. Principal component analysis was performed on the filtered exposure matrix using a truncated singular value decomposition approach into 20 principal components. Similar analyses were done for primary-suspect drugs (15 PC with minimum of 50 patients), indications (20 PC, minimum of 100 patients), and outcomes (15 PC, minimum of 50 patients). DrugPCs summarized latent patterns of correlated drug exposure and were included in multivariable models alongside total medication burden (NumDrugs) to adjust for structured polypharmacy.

We constructed logistic regression models over the FAERS dataset to evaluate the association between HIV and SCAR outcomes. Three nested models were specified:

1. Unadjusted model including HIV status only
2. Demographic-adjusted model including age (modeled using natural splines with 4 degrees of freedom to account for nonlinearity), sex, geographic region, and report year
3. Fully adjusted model additionally including polypharmacy (NumDrugs) and drug exposure principal components (DrugPC_*).

### Interaction Analysis

To assess effect modification by HIV, we constructed logistic regression models including terms for drug exposure (PS), HIV status, and their interaction (Drug × HIV) over the entire dataset. For report *i*:

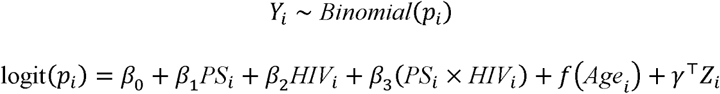

Where:

*Y_l_* : SCAR outcome (Any SCAR / SJS-TEN / DRESS)
PS_*i*_ : drug is primary suspect
HIV_*i*_: if HIV+ (post-imputation)
*f(Age_i_)*: spline for age
*Z_i_* :sex, region, year, number of drugs

Analyses were restricted to strata with a minimum of 5 events in the 2 x 2 contingency table of HIV-pos/neg and drug exposure positive/negative (for each drug) to ensure stability of estimates and avoid sparse-data bias. These models estimated drug-associated SCAR risk separately in HIV-positive (OR = exp(β_1_+β_3_)) and HIV-negative (OR = exp(β_1_)) populations.

### Ethical Considerations

This study used publicly available, de-identified data and was exempt from institutional review board oversight.

## Results

### HIV Classification and Cohort Assembly

Among 13,986,839 FAERS reports submitted between 2013 and 2023, 153,203 had exposure to at least one antiretroviral medication. HIV status was assigned using a hierarchical classification strategy integrating antiretroviral exposure, indication text, and machine-learning imputation (**Figure S1**). Among antiretroviral-exposed reports, 50,021 had ambiguous indications and underwent random forest imputation. The classifier demonstrated excellent out-of-bag performance (PFC = 0.00097). Overall, 139,539 reports (1.0%) were classified as HIV-positive. Among these, 93,449 had an antiretroviral medication with an HIV-positive indication (66.97%), 40,172 had antiretroviral exposure with ambiguous indications and were classified as HIV-positive by imputation (28.79%), 5,869 had explicit HIV evidence without recorded antiretroviral exposure (4.21%), and 49 had antiretroviral exposure with HIV status rescued by explicit HIV terms elsewhere in the report (0.04%).

### Multiple Antiretroviral and Non-Antiretroviral Medications Contribute to HIV-Associated SCAR

Demographic and clinical characteristics of HIV-positive SCAR reports are summarized in **Table 1**. Among 139,539 HIV-positive patients, 899 presented with at least one SCAR, with 440 presenting with SJS/TEN, 414 with DRESS, 65 with AGEP, and 1 with GBFDE. Median age was 48 years in both the SCAR and non-SCAR cohorts. Among all HIV-positive patients presenting with SCAR, 11.9% died (n = 107), and specifically among SJS/TEN patients 20% died (n = 88) compared to 7.1% of all HIV-positive patients.

**Table 1.** Demographic and clinical characteristics of severe cutaneous adverse reaction (SCAR) reports in HIV-positive patients.

| Demographic Summary of HIV-Positive Patients by SCAR Diagnosis |  |  |  |  |  |  |  |
| --- | --- | --- | --- | --- | --- | --- | --- |
| Characteristic | HIV+ Total | No SCAR | Total SCAR | SJS/TEN | DRESS | AGEP | GBFDE |
| Total N | 139539 | 138640 | 899 | 440 | 414 | 65 | 1 |
| Age, median [IQR] | 48 [35–57] | 48 [35–57] | 45 [32.5–56] | 40 [28–53] | 49 [40–57] | 58 [45–73] | 69 [69–69] |
| Sex | Female: 36413 (26.1%)<br>Male: 77467 (55.5%)<br>Not Specified: 25659 (18.4%) | Female: 36058 (26%)<br>Male: 77117 (55.6%)<br>Not Specified: 25465 (18.4%) | Female: 355 (39.5%)<br>Male: 350 (38.9%)<br>Not Specified: 194 (21.6%) | Female: 168 (38.2%)<br>Male: 154 (35%)<br>Not Specified: 118 (26.8%) | Female: 180 (43.5%)<br>Male: 187 (45.2%)<br>Not Specified: 47 (11.4%) | Female: 10 (15.4%)<br>Male: 23 (35.4%)<br>Not Specified: 32 (49.2%) | Not Specified: 1 (100%) |
| Region | East Asia & Pacific: 7479 (5.4%)<br>Europe & Central Asia: 35239 (25.3%)<br>Latin America & Caribbean: 2810 (2%)<br>Middle East & North Africa: 820 (0.6%)<br>North America: 83838 (60.1%)<br>South Asia: 1084 (0.8%)<br>Sub-Saharan Africa: 8173 (5.9%)<br>Unknown: 96 (0.1%) | East Asia & Pacific: 7410 (5.3%)<br>Europe & Central Asia: 34819 (25.1%)<br>Latin America & Caribbean: 2784 (2%)<br>Middle East & North Africa: 816 (0.6%)<br>North America: 83689 (60.4%)<br>South Asia: 997 (0.7%)<br>Sub-Saharan Africa: 8029 (5.8%)<br>Unknown: 96 (0.1%) | East Asia & Pacific: 69 (7.7%)<br>Europe & Central Asia: 420 (46.7%)<br>Latin America & Caribbean: 26 (2.9%)<br>Middle East & North Africa: 4 (0.4%)<br>North America: 149 (16.6%)<br>South Asia: 87 (9.7%)<br>Sub-Saharan Africa: 144 (16%) | East Asia & Pacific: 36 (8.2%)<br>Europe & Central Asia: 96 (21.8%)<br>Latin America & Caribbean: 17 (3.9%)<br>Middle East & North Africa: 1 (0.2%)<br>North America: 86 (19.5%)<br>South Asia: 66 (15%)<br>Sub-Saharan Africa: 138 (31.4%) | East Asia & Pacific: 32 (7.7%)<br>Europe & Central Asia: 284 (68.6%)<br>Latin America & Caribbean: 9 (2.2%)<br>North America: 63 (15.2%)<br>South Asia: 20 (4.8%)<br>Sub-Saharan Africa: 6 (1.4%) | East Asia & Pacific: 2 (3.1%)<br>Europe & Central Asia: 56 (86.2%)<br>Middle East & North Africa: 3 (4.6%)<br>North America: 3 (4.6%)<br>South Asia: 1 (1.5%) | North America: 1 (100%) |
| Outcome | Alive: 129686 (92.9%)<br>Dead: 9853 (7.1%) | Alive: 128894 (93%)<br>Dead: 9746 (7%) | Alive: 792 (88.1%)<br>Dead: 107 (11.9%) | Alive: 352 (80%)<br>Dead: 88 (20%) | Alive: 385 (93%)<br>Dead: 29 (7%) | Alive: 64 (98.5%)<br>Dead: 1 (1.5%) | Alive: 1 (100%) |

Among HIV-positive SCAR reports, 116 distinct primary suspect drugs were identified (**Table S2**). Antiretroviral medications accounted for a substantial proportion of reports, particularly nevirapine (n=175), lamivudine (n=85), abacavir (n=62), raltegravir (n=48), and emtricitabine (n=47). However, numerous non-antiretroviral medications were also represented, including hydroxychloroquine (n=29), trimethoprim (n=25), sulfamethoxazole (n=24), azithromycin (n=12), ribavirin (n=11), and rifampin (n=9). Distinct patterns were observed across SCAR phenotypes. SJS/TEN reports were dominated by nevirapine and several nucleoside reverse transcriptase inhibitors, whereas DRESS reports were frequently associated with abacavir, raltegravir, lamivudine, and rifampin. AGEP demonstrated a different exposure profile, with hydroxychloroquine accounting for the largest number of reports. Collectively, these findings demonstrate that SCAR among PLWH occurs across a diverse medication landscape extending well beyond antiretroviral therapy alone.

To determine whether specific medications were disproportionately associated with SCAR among PLWH, we next performed drug-level disproportionality analyses using primary suspect designations (**Figure 3**). Several established SCAR-associated medications demonstrated elevated reporting odds ratios across multiple phenotypes. Among antiretroviral agents, nevirapine exhibited the strongest associations with both any SCAR and SJS/TEN, while raltegravir demonstrated prominent associations with DRESS. Among non-antiretroviral medications, trimethoprim, sulfamethoxazole, carbamazepine, clarithromycin, levetiracetam, rifampin, and lansoprazole were associated with increased reporting across one or more SCAR phenotypes.

**Figure 2.**
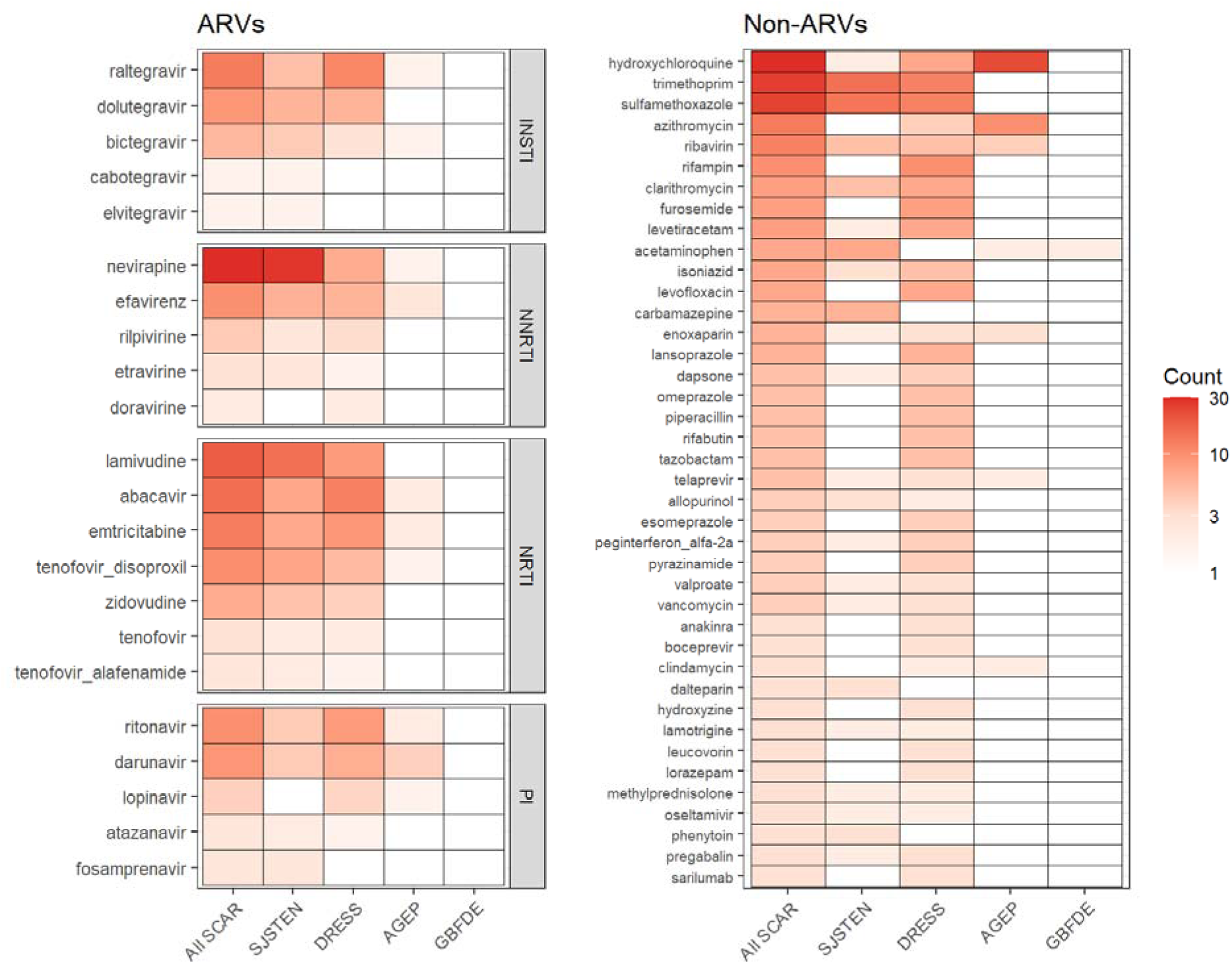
Primary suspect drugs reported among HIV-positive individuals with severe cutaneous adverse reactions. Heatmap showing the number of HIV-positive reports in which each medication was designated as the primary suspect (PS) drug for SCAR, stratified by reaction phenotype. Antiretroviral agents are grouped by drug class (integrase strand transfer inhibitors [INSTIs], non-nucleoside reverse transcriptase inhibitors [NNRTIs], nucleoside reverse transcriptase inhibitors [NRTIs], and protease inhibitors [PIs]). Non-antiretroviral medications include established SCAR-associated antibiotics, anticonvulsants, proton pump inhibitors, and other agents. Cell shading represents the number of HIV-positive reports for each drug-phenotype combination.

**Figure 3.**
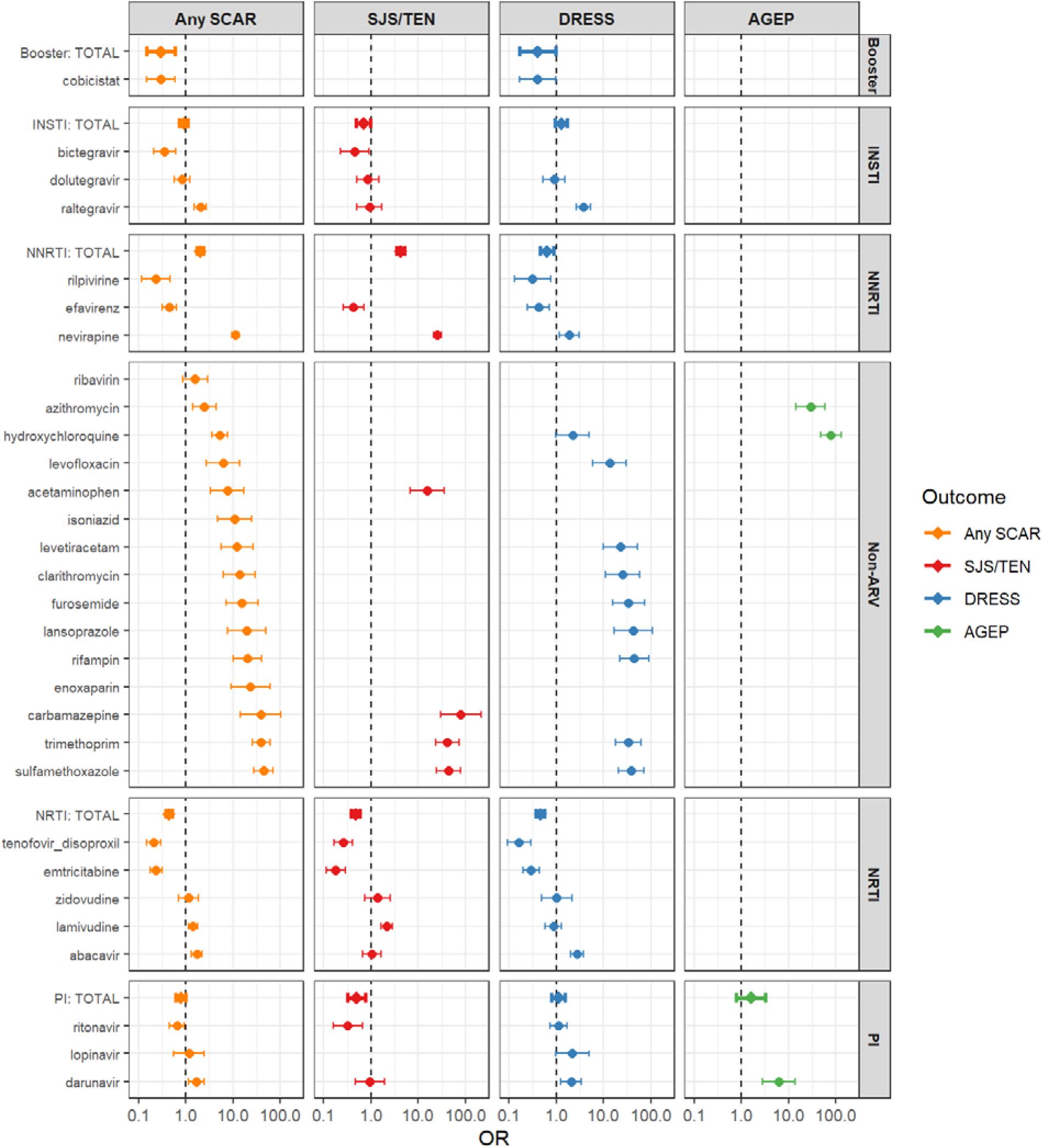
Primary suspect drug reporting odds ratios (RORs) for SCAR among HIV-positive individuals. Forest plots display disproportionality estimates for selected medications with at least five exposed cases. RORs greater than 1 indicate disproportionate reporting relative to HIV-negative reports.

### Demographic and Medication Exposure Explain Much of the HIV-SCAR Association

We next evaluated the association between HIV infection and SCAR using sequential logistic regression models incorporating demographic and drug exposure covariates. In unadjusted analyses, HIV infection was strongly associated with all SCAR outcomes (**Figure S4**). HIV-positive status was associated with increased odds of any SCAR (OR 2.26, 95% CI [2.11-2.41]), SJS/TEN (OR 2.57 [2.33-2.82]), and DRESS (OR 2.23 [2.46-2.82]). Adjustment for demographic factors, including age, sex, geographic region, and calendar year, substantially attenuated these associations (**Figure S5**). Following demographic adjustment, the odds ratios decreased to 1.37 (95% CI 1.28-1.47) for any SCAR, 1.55 (1.41-1.71) for SJS/TEN, and 1.32 (1.20-1.46) for DRESS, indicating that demographic differences accounted for a substantial proportion of the observed HIV-associated risk.

We next incorporated polypharmacy burden and principal components representing structured drug exposure into the regression models. Principal component analysis demonstrated substantial underlying structure within drug exposure data, with approximately 8-14 components explaining 80% of total variance across exposure matrices (**Figure S3**). Inclusion of these variables resulted in further attenuation of HIV-associated risk (**Figure 4**). In the fully adjusted model, HIV remained associated with any SCAR (OR 1.12, 95% CI [1.04-1.20]) and SJS/TEN (OR 1.31 [1.19-1.45]), whereas the association with DRESS was no longer significant (OR 1.02 [0.92-1.13]). Comparison of effect estimates across sequential models demonstrated that both demographic characteristics and medication exposure contributed substantially to the observed HIV-SCAR association (**Figure 5**). Drug exposure adjustment produced the largest attenuation for DRESS, whereas a modest residual association persisted for SJS/TEN despite extensive adjustment.

**Figure 4.**
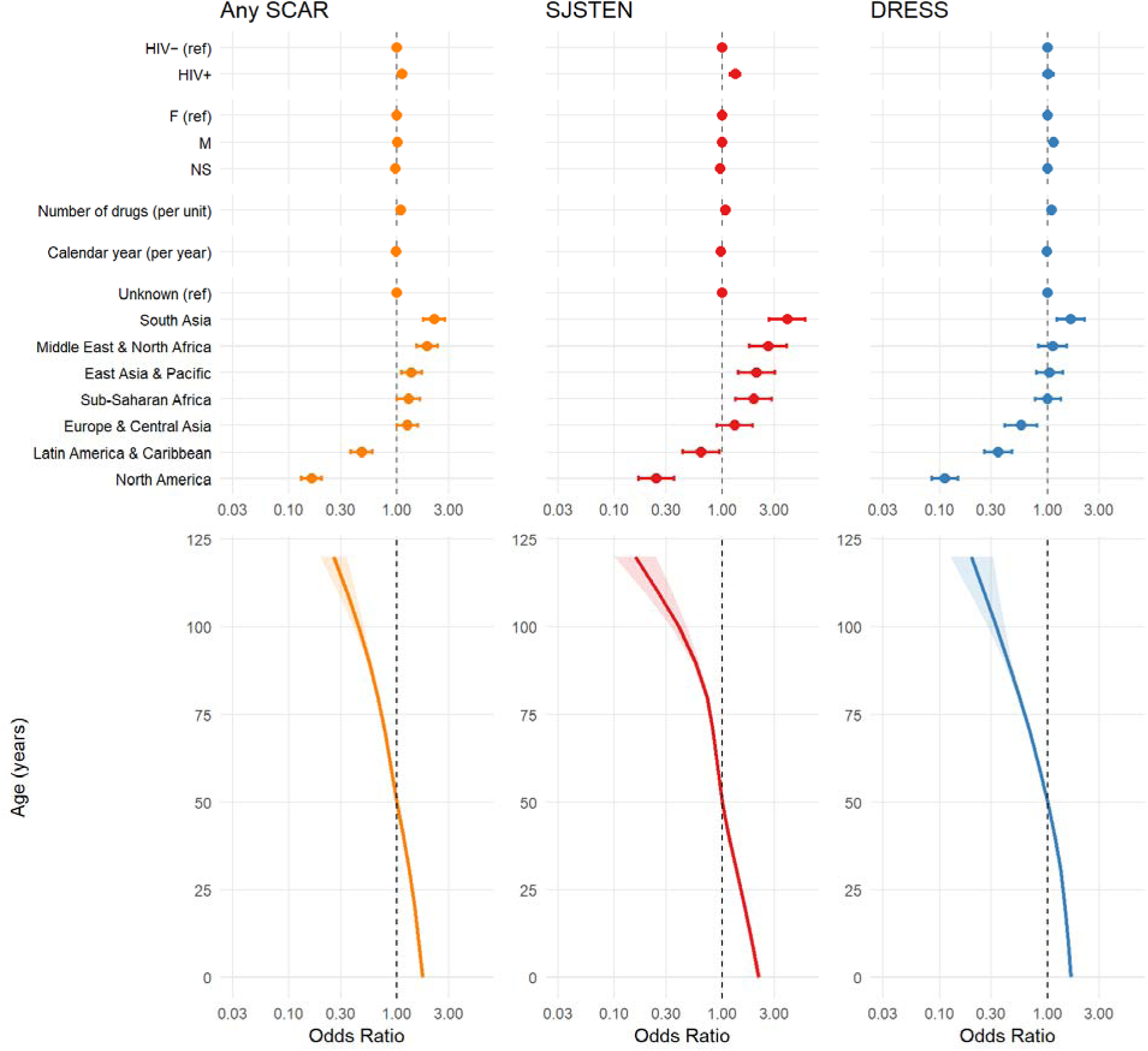
Fully adjusted logistic regression models evaluating the association between HIV infection and SCAR outcomes. Models adjusted for age (natural cubic spline with 4 degrees of freedom), sex, geographic region, reporting year, medication burden, and drug principal components representing structured medication exposure. HIV infection was associated with increased odds of any SCAR (OR 1.12, 95% CI [1.04-1.20]), SJS/TEN (OR 1.31 [1.19-1.45]), but not DRESS (OR 1.02 [0.92-1.13]).

**Figure 5.**
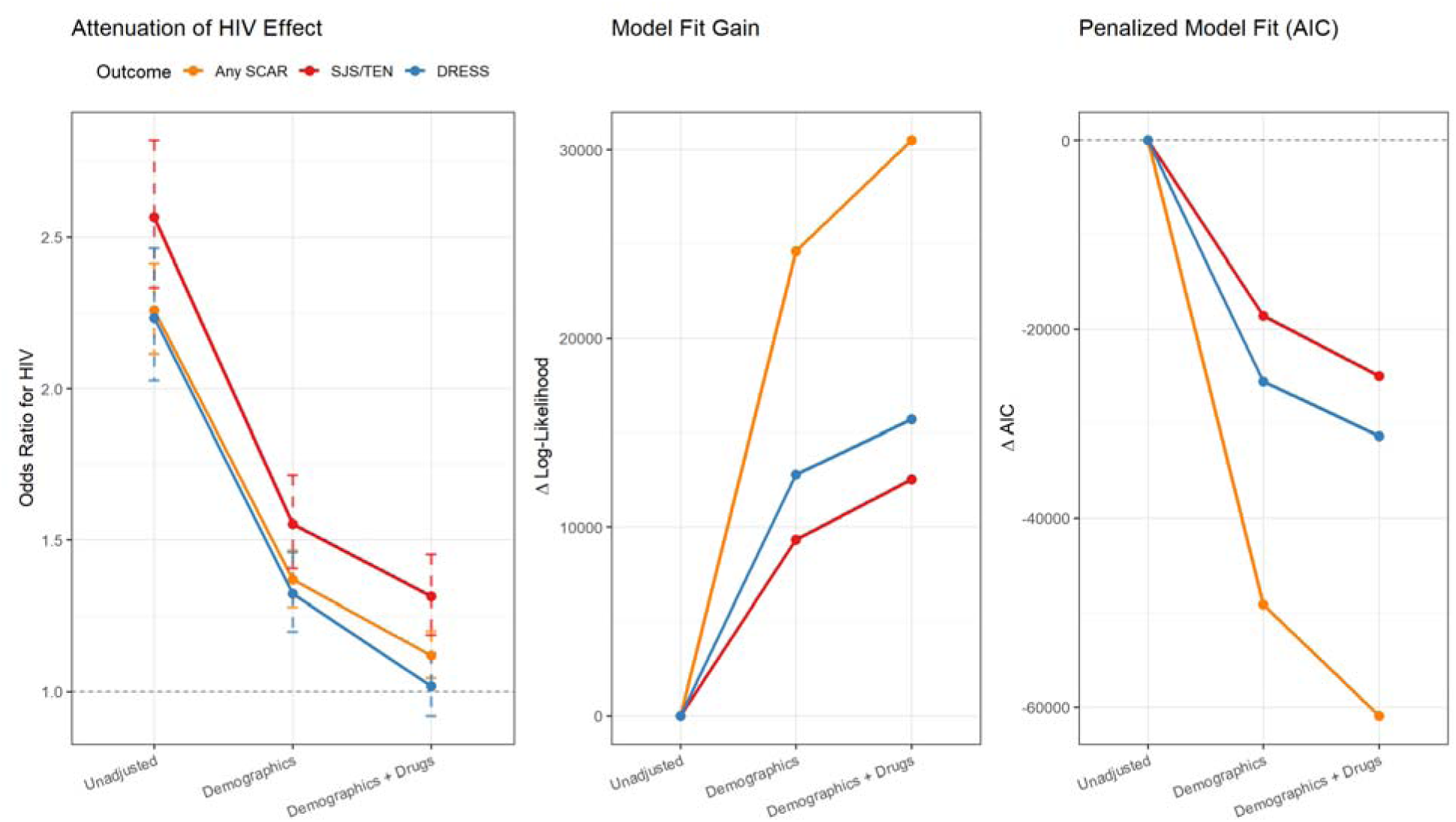
Attenuation of HIV-associated SCAR risk across sequential regression models. Odds ratios for HIV are shown before adjustment, after demographic adjustment, and after adjustment for structured medication exposure. Model fit statistics demonstrate incremental explanatory value of demographic and drug exposure variables.

To determine whether HIV modifies susceptibility to specific drug triggers, we constructed drug-by-HIV interaction models using logistic regression with an interaction term between each PS drug and HIV. Although most medications demonstrated broadly similar effect sizes in HIV-positive and HIV-negative individuals (**Figure S6**), several drugs exhibited evidence of selective amplification among PLWH (**Figure 6**). Notable examples included carbamazepine and clarithromycin for SJS/TEN, and levetiracetam, lansoprazole, and furosemide for DRESS.

**Figure 6.**
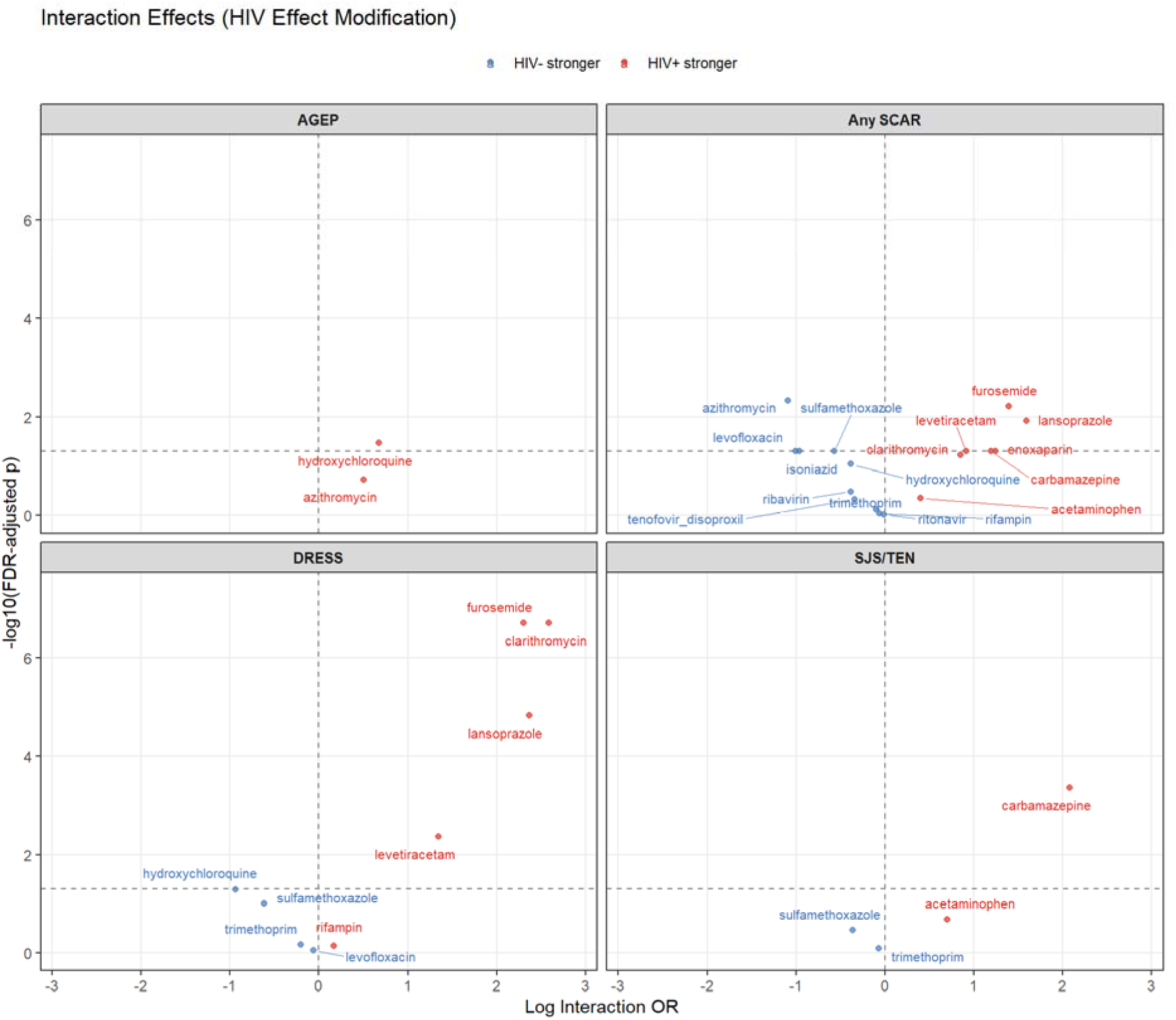
Interaction between HIV infection and primary suspect drugs for SCAR outcomes. Interaction odds ratios represent multiplicative effect modification, with values greater than 1 indicating stronger drug-associated SCAR reporting among HIV-positive individuals. Selective amplification was observed for several medications, whereas others demonstrated little evidence of HIV-specific modification.

As a final analysis, we analyzed co-exposure across causative agents (**Table S7**). Across HIV-positive SCAR reports, co-exposure to at least one additional drug with an ROR025 greater than 1 was extremely common for most non-ARV primary suspect drugs. For overall SCAR, every case involving carbamazepine, clarithromycin, rifampin, hydroxychloroquine, levetiracetam, lansoprazole, or furosemide had at least one such co-medication, compared with 56% of TMP-SMX cases. The same pattern largely persisted for SJS/TEN and DRESS, although TMP-SMX was less consistently accompanied by another ROR025-positive drug, particularly in DRESS, where only 2 of 11 cases (18.2%) had one. Recurrent co-medications included antiretrovirals such as nevirapine, lamivudine, abacavir, raltegravir, ritonavir, and darunavir, as well as multidrug tuberculosis regimens and other known SCAR-associated agents, suggesting substantial competing causality and treatment-context confounding for many of these signals.

## Discussion

In this large pharmacovigilance study of nearly 14 million FAERS reports, HIV infection was associated with increased reporting of SCAR across multiple phenotypes. However, this association was substantially attenuated after adjustment for demographic characteristics and structured medication exposure. Following full adjustment, only modest associations remained for any SCAR and SJS/TEN, while the association with DRESS was no longer significant. Furthermore, HIV did not uniformly amplify drug-associated risk. Rather, HIV selectively increased risk for a limited number of drug-SCAR combinations. Together, these findings suggest that the relationship between HIV and SCAR reflects both differential drug exposure and biologic susceptibility, with the latter acting in a drug-specific rather than global manner.

Our findings are consistent with prior literature suggesting that people living with HIV experience higher rates of drug hypersensitivity than the general population.^8–12^ A widely-cited cohort study states that the incidence of SJS/TEN in HIV-positive patients is 100 times that of the background population, at 1-2 per 1000, though the biological basis and risk factors leading to this increase are incompletely defined.^13^ Multiple mechanisms have been proposed, including chronic immune activation, depletion of CD4+ regulatory T cells in skin, viral co-infections, oxidative stress, and changes in drug metabolism.^14–16^ However, HIV-positive individuals also experience markedly different medication exposures than the general population, including antiretroviral therapy, treatment of opportunistic infections, and increased polypharmacy. Consequently, it has remained difficult to determine the extent to which HIV itself contributes to risk versus the medications commonly prescribed within this population.

Our findings suggest that differential medication exposure and demographic factors account for a substantial portion of the observed HIV-SCAR association. The magnitude of attenuation observed after adjustment for structured drug exposure was striking. At the same time, complete attenuation was not observed. HIV remained independently associated with SJS/TEN after extensive adjustment, and several drugs demonstrated stronger associations among PLWH than among HIV-negative individuals. These findings argue against a purely exposure-driven explanation and support a role for host susceptibility. Importantly, however, this susceptibility appeared highly selective. Most drugs demonstrated remarkably similar effect sizes across HIV strata, whereas only limited number exhibited evidence of amplification. This pattern is inconsistent with a model in which HIV broadly increases SCAR risk across all medications and instead supports a model in which HIV modifies susceptibility to specific drug triggers. These observations align with contemporary concepts of delayed-type drug hypersensitivity.^17,18^ Within this framework, HIV may function as a susceptibility state that lowers the threshold for clinically apparent hypersensitivity only in selected biologic contexts. The preferential amplification observed for drugs such as carbamazepine and clarithromycin suggests that HIV-related immune dysregulation may interact with specific pathways of antigen presentation or T-cell activation rather than producing a generalized increase in immune reactivity.

The co-medication analysis further underscored the importance of treatment context. Most non-antiretroviral primary suspect drugs were reported alongside at least one additional medication with an ROR025 greater than 1, and several signals occurred within highly complex regimens involving antiretroviral, antituberculosis, antimicrobial, or other recognized SCAR-associated drugs. This was particularly evident for clarithromycin, rifampin, hydroxychloroquine, levetiracetam, and furosemide, whereas TMP-SMX more often appeared in relatively simpler regimens. These findings reinforce the likelihood of substantial competing causality and confounding by indication in many HIV-associated SCAR reports.

The differential behavior of DRESS and SJS/TEN is particularly noteworthy. For DRESS, the association with HIV was almost entirely explained by demographics and medication use. These findings indicate that a meaningful fraction of the historical HIV-SCAR signal likely reflects differences in exposure to high-risk medications rather than a uniform increase in biologic susceptibility. Although the present study cannot establish mechanism, these findings raise the possibility that HIV-related susceptibility differs across SCAR phenotypes. Future studies integrating HLA genotype, viral reactivation status, and immune profiling may help clarify whether distinct pathways underlie these observations.

From a clinical perspective, our findings suggest that HIV should not be viewed as conferring uniformly elevated risk for severe drug hypersensitivity. Instead, excess risk appears concentrated within specific drug–host combinations. This distinction has important implications for risk assessment and prescribing practices. Rather than assuming heightened susceptibility across all medications, clinicians may be better served by focusing on specific high-risk drugs and clinical contexts in which HIV appears to amplify risk.

This study has several limitations. FAERS is a spontaneous reporting database and cannot establish incidence, causality, or absolute risk. Residual confounding remains possible despite adjustment for demographic factors, polypharmacy, and structured medication exposure. HIV status was inferred using medication and indication data for a subset of reports, although classifier performance was excellent. Finally, differential reporting behavior and healthcare utilization may influence observed associations. Despite these limitations, the scale of FAERS enabled comprehensive evaluation of HIV-associated SCAR across multiple phenotypes and medication classes. By explicitly accounting for structured drug exposure, we demonstrate that much of the HIV-SCAR association is attributable to differences in medication use, while the residual signal is concentrated within specific drugs and phenotypes. These findings support a model in which HIV functions as a context-dependent host susceptibility state rather than a universal amplifier of severe drug hypersensitivity.

## Data Availability

All data produced in the present study are available upon reasonable request to the authors

## Abbreviation

SCAR: Severe Cutaneous Adverse Reactions
SJS/TEN: Stevens-Johnson Syndrome/Toxic Epidermal Necrolysis
DRESS: Drug Reaction with Eosinophilia and Systemic Symptoms
AGEP: Acute Generalized Exanthematous Pustulosis
GBFDE: Generalized Bullous Fixed Drug Eruption
FAERS: FDA Adverse Event Reporting System
ROR: Reporting Odds Ratio
ARV: Antiretroviral
PLWH: Person Living with HIV
NRTI: Nucleoside Reverse Transcriptase Inhibitor
NNRTI: Non-Nucleoside Reverse Transcriptase Inhibitor
PI: Protease Inhibitor
INSTI: Integrase Strand Transfer Inhibitor

## Contributions

EMM conceived the project, wrote the manuscript, and conducted all analyses. DP assisted with analyses and edited the manuscript. EP, AA, MMP, CAS, and MSK assisted with analyses and reviewed the manuscript.

## Acknowledgements

EJP is supported by the following grants from the National Institutes of Health (NIH): NIH U01AI154659, NIH P50GM115305, NIH R01HG010863, NIH R21AI139021, NIH R01AI152183, and NIH 2 D43 TW010559. EJP is also supported by the National Health and Medical Research Council of Australia. EMM is funded by a Vanderbilt University Medical Center internal career development award (Vanderbilt Faculty Research Scholars).

## AI Use Statement

GPT4o and GPT5 were used to optimize and debug code and assist with manuscript editing. Nano Banana 2 was used to generate Figure S1 (with manual editing afterwards). The authors take responsibility for all code and data generated.

---

**Figure S1.**
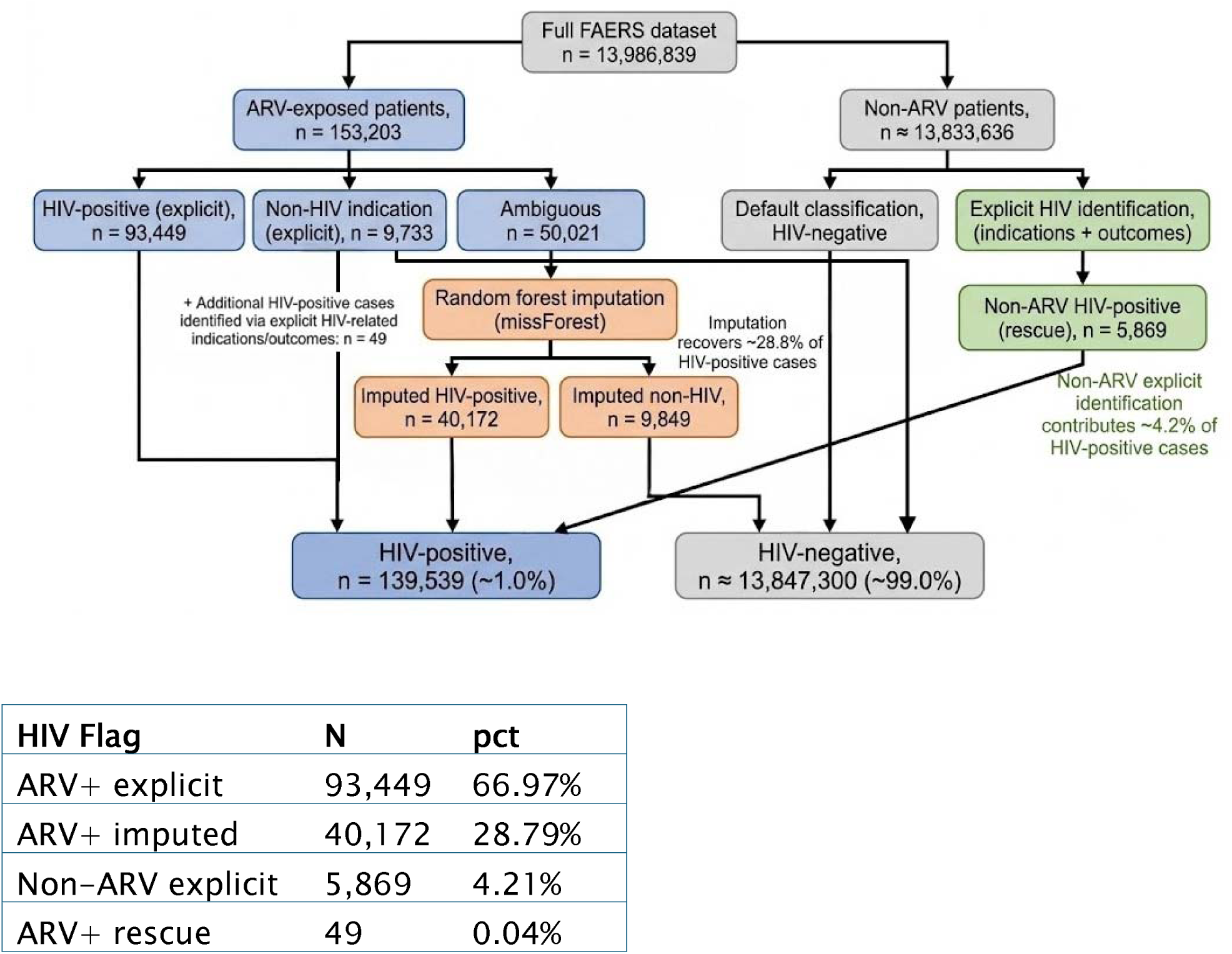
HIV status classification and imputation pipeline. Hierarchical workflow used to assign HIV status within the FAERS database. Among antiretroviral (ARV)-exposed reports, individuals with explicit HIV-related indications were classified as HIV-positive, whereas those with explicit non-HIV indications were classified as HIV-negative. Reports with ambiguous indications underwent random forest imputation using demographic characteristics, geographic region, medication burden, clinical outcomes, antiretroviral exposures, and drug principal components. Among non-ARV-exposed reports, HIV-positive cases were identified through explicit HIV-related indications or outcomes. The random forest model demonstrated excellent out-of-bag performance (proportion falsely classified [PFC] = 0.00097), indicating minimal classification error. Application of this framework yielded 139,539 HIV-positive reports (approximately 1.0% of all FAERS reports), including 40,172 HIV-positive reports identified through imputation and 5,869 additional HIV-positive reports identified among non-ARV-exposed individuals.

**Table S2.** Primary-Suspect Drugs in SCAR among PLWH.

| Group | drug_name | Total SCAR | SJS-TEN | DRESS | AGEP | GBFDE |
| --- | --- | --- | --- | --- | --- | --- |
| TOTAL | Any Drug | 930 | 458 | 425 | 67 | 1 |
| NRTI | NRTI TOTAL | 251 | 131 | 118 | 5 | 0 |
| NNRTI | NNRTI TOTAL | 221 | 178 | 39 | 4 | 0 |
| PI | PI TOTAL | 73 | 21 | 48 | 10 | 0 |
| INSTI | INSTI TOTAL | 91 | 35 | 57 | 2 | 0 |
| Booster | Booster TOTAL | 8 | 4 | 5 | 0 | 0 |
| Non-ARV | Non-ARV TOTAL | 286 | 89 | 158 | 46 | 1 |
| NRTI | lamivudine | 85 | 60 | 25 | 0 | 0 |
| NRTI | abacavir | 62 | 19 | 44 | 2 | 0 |
| NRTI | emtricitabine | 47 | 18 | 27 | 2 | 0 |
| NRTI | tenofovir_disoproxil | 33 | 20 | 12 | 1 | 0 |
| NRTI | zidovudine | 17 | 10 | 7 | 0 | 0 |
| NRTI | tenofovir | 4 | 2 | 2 | 0 | 0 |
| NRTI | tenofovir_alafenamide | 3 | 2 | 1 | 0 | 0 |
| NNRTI | nevirapine | 175 | 157 | 17 | 1 | 0 |
| NNRTI | efavirenz | 32 | 15 | 14 | 3 | 0 |
| NNRTI | rilpivirine | 8 | 3 | 5 | 0 | 0 |
| NNRTI | etravirine | 4 | 3 | 1 | 0 | 0 |
| NNRTI | doravirine | 2 | 0 | 2 | 0 | 0 |
| PI | ritonavir | 32 | 8 | 25 | 2 | 0 |
| PI | darunavir | 28 | 8 | 16 | 7 | 0 |
| PI | lopinavir | 7 | 0 | 6 | 1 | 0 |
| PI | atazanavir | 3 | 2 | 1 | 0 | 0 |
| PI | fosamprenavir | 3 | 3 | 0 | 0 | 0 |
| INSTI | raltegravir | 48 | 11 | 39 | 1 | 0 |
| INSTI | dolutegravir | 28 | 14 | 14 | 0 | 0 |
| INSTI | bictegravir | 13 | 8 | 4 | 1 | 0 |
| INSTI | cabotegravir | 1 | 1 | 0 | 0 | 0 |
| INSTI | elvitegravir | 1 | 1 | 0 | 0 | 0 |
| Booster | cobicistat | 8 | 4 | 5 | 0 | 0 |
| Non-ARV | hydroxychloroquine | 29 | 1 | 6 | 22 | 0 |
| Non-ARV | trimethoprim | 25 | 14 | 11 | 0 | 0 |
| Non-ARV | sulfamethoxazole | 24 | 13 | 11 | 0 | 0 |
| Non-ARV | azithromycin | 12 | 0 | 3 | 9 | 0 |
| Non-ARV | ribavirin | 11 | 4 | 4 | 3 | 0 |
| Non-ARV | rifampin | 9 | 0 | 9 | 0 | 0 |
| Non-ARV | clarithromycin | 7 | 4 | 6 | 0 | 0 |
| Non-ARV | furosemide | 7 | 0 | 7 | 0 | 0 |
| Non-ARV | levetiracetam | 7 | 1 | 6 | 0 | 0 |
| Non-ARV | acetaminophen | 6 | 6 | 0 | 1 | 1 |
| Non-ARV | isoniazid | 6 | 2 | 4 | 0 | 0 |
| Non-ARV | levofloxacin | 6 | 0 | 6 | 0 | 0 |
| Non-ARV | carbamazepine | 5 | 5 | 0 | 0 | 0 |
| Non-ARV | enoxaparin | 5 | 1 | 2 | 2 | 0 |
| Non-ARV | lansoprazole | 5 | 0 | 5 | 0 | 0 |
| Non-ARV | dapsone | 4 | 1 | 3 | 0 | 0 |
| Non-ARV | omeprazole | 4 | 0 | 4 | 0 | 0 |
| Non-ARV | piperacillin | 4 | 0 | 4 | 0 | 0 |
| Non-ARV | rifabutin | 4 | 0 | 4 | 0 | 0 |
| Non-ARV | tazobactam | 4 | 0 | 4 | 0 | 0 |
| Non-ARV | telaprevir | 4 | 1 | 2 | 1 | 0 |
| Non-ARV | allopurinol | 3 | 2 | 1 | 0 | 0 |
| Non-ARV | esomeprazole | 3 | 0 | 3 | 0 | 0 |
| Non-ARV | peginterferon_alfa-2a | 3 | 1 | 3 | 0 | 0 |
| Non-ARV | pyrazinamide | 3 | 0 | 3 | 0 | 0 |
| Non-ARV | valproate | 3 | 1 | 2 | 0 | 0 |
| Non-ARV | vancomycin | 3 | 1 | 2 | 0 | 0 |
| Non-ARV | anakinra | 2 | 0 | 2 | 0 | 0 |
| Non-ARV | boceprevir | 2 | 0 | 2 | 0 | 0 |
| Non-ARV | clindamycin | 2 | 0 | 1 | 1 | 0 |
| Non-ARV | dalteparin | 2 | 2 | 0 | 0 | 0 |
| Non-ARV | hydroxyzine | 2 | 0 | 2 | 0 | 0 |
| Non-ARV | lamotrigine | 2 | 1 | 1 | 0 | 0 |
| Non-ARV | leucovorin | 2 | 0 | 2 | 0 | 0 |
| Non-ARV | lorazepam | 2 | 0 | 2 | 0 | 0 |
| Non-ARV | methylprednisolone | 2 | 1 | 1 | 0 | 0 |
| Non-ARV | oseltamivir | 2 | 1 | 1 | 0 | 0 |
| Non-ARV | phenytoin | 2 | 2 | 0 | 0 | 0 |
| Non-ARV | pregabalin | 2 | 1 | 2 | 0 | 0 |
| Non-ARV | sarilumab | 2 | 0 | 2 | 0 | 0 |
| Non-ARV | sulfasalazine | 2 | 0 | 2 | 0 | 0 |
| Non-ARV | acetazolamide | 1 | 0 | 1 | 0 | 0 |
| Non-ARV | acetylcysteine | 1 | 0 | 1 | 0 | 0 |
| Non-ARV | acyclovir | 1 | 1 | 0 | 0 | 0 |
| Non-ARV | amiodarone | 1 | 0 | 1 | 0 | 0 |
| Non-ARV | amlodipine | 1 | 0 | 0 | 1 | 0 |
| Non-ARV | amoxicillin | 1 | 0 | 1 | 0 | 0 |
| Non-ARV | aspirin | 1 | 1 | 0 | 0 | 0 |
| Non-ARV | bupropion | 1 | 1 | 0 | 0 | 0 |
| Non-ARV | cefixime | 1 | 1 | 0 | 0 | 0 |
| Non-ARV | ceftriaxone | 1 | 0 | 0 | 1 | 0 |
| Non-ARV | chloroquine | 1 | 0 | 0 | 1 | 0 |
| Non-ARV | ciprofloxacin | 1 | 0 | 1 | 0 | 0 |
| Non-ARV | cisplatin | 1 | 0 | 0 | 1 | 0 |
| Non-ARV | clofazimine | 1 | 0 | 1 | 0 | 0 |
| Non-ARV | clonazepam | 1 | 1 | 0 | 0 | 0 |
| Non-ARV | clonidine | 1 | 0 | 1 | 0 | 0 |
| Non-ARV | clopidogrel | 1 | 0 | 1 | 0 | 0 |
| Non-ARV | clozapine | 1 | 0 | 1 | 0 | 0 |
| Non-ARV | colchicine | 1 | 1 | 0 | 0 | 0 |
| Non-ARV | daptomycin | 1 | 0 | 1 | 0 | 0 |
| Non-ARV | elbasvir | 1 | 1 | 0 | 0 | 0 |
| Non-ARV | ethambutol | 1 | 0 | 1 | 0 | 0 |
| Non-ARV | fluconazole | 1 | 1 | 0 | 0 | 0 |
| Non-ARV | gabapentin | 1 | 1 | 0 | 0 | 0 |
| Non-ARV | interferon_alfa-2a | 1 | 0 | 0 | 1 | 0 |
| Non-ARV | iopamidol | 1 | 0 | 1 | 0 | 0 |
| Non-ARV | iopromide | 1 | 0 | 1 | 0 | 0 |
| Non-ARV | itraconazole | 1 | 1 | 1 | 0 | 0 |
| Non-ARV | ivermectin | 1 | 1 | 0 | 0 | 0 |
| Non-ARV | ketoconazole | 1 | 1 | 0 | 0 | 0 |
| Non-ARV | lacosamide | 1 | 1 | 0 | 0 | 0 |
| Non-ARV | loperamide | 1 | 1 | 0 | 0 | 0 |
| Non-ARV | lumefantrine | 1 | 1 | 0 | 0 | 0 |
| Non-ARV | methotrexate | 1 | 1 | 0 | 0 | 0 |
| Non-ARV | metronidazole | 1 | 1 | 0 | 0 | 0 |
| Non-ARV | nitrofurantoin | 1 | 0 | 1 | 0 | 0 |
| Non-ARV | nystatin | 1 | 0 | 1 | 0 | 0 |
| Non-ARV | ondansetron | 1 | 0 | 0 | 1 | 0 |
| Non-ARV | oxycodone | 1 | 1 | 0 | 0 | 0 |
| Non-ARV | pantoprazole | 1 | 0 | 1 | 0 | 0 |
| Non-ARV | paroxetine | 1 | 1 | 0 | 0 | 0 |
| Non-ARV | pentoxifylline | 1 | 0 | 1 | 0 | 0 |
| Non-ARV | prednisolone | 1 | 0 | 0 | 1 | 0 |
| Non-ARV | prednisone | 1 | 1 | 0 | 0 | 0 |
| Non-ARV | risperidone | 1 | 0 | 1 | 0 | 0 |
| Non-ARV | rituximab | 1 | 1 | 0 | 0 | 0 |
| Non-ARV | testosterone | 1 | 1 | 0 | 0 | 0 |
| Non-ARV | thiamine | 1 | 0 | 1 | 0 | 0 |
| Non-ARV | tigecycline | 1 | 0 | 1 | 0 | 0 |
| Non-ARV | ustekinumab | 1 | 1 | 0 | 0 | 0 |
| Non-ARV | valacyclovir | 1 | 0 | 1 | 0 | 0 |
| Non-ARV | valganciclovir | 1 | 0 | 1 | 0 | 0 |

**Figure S3.**
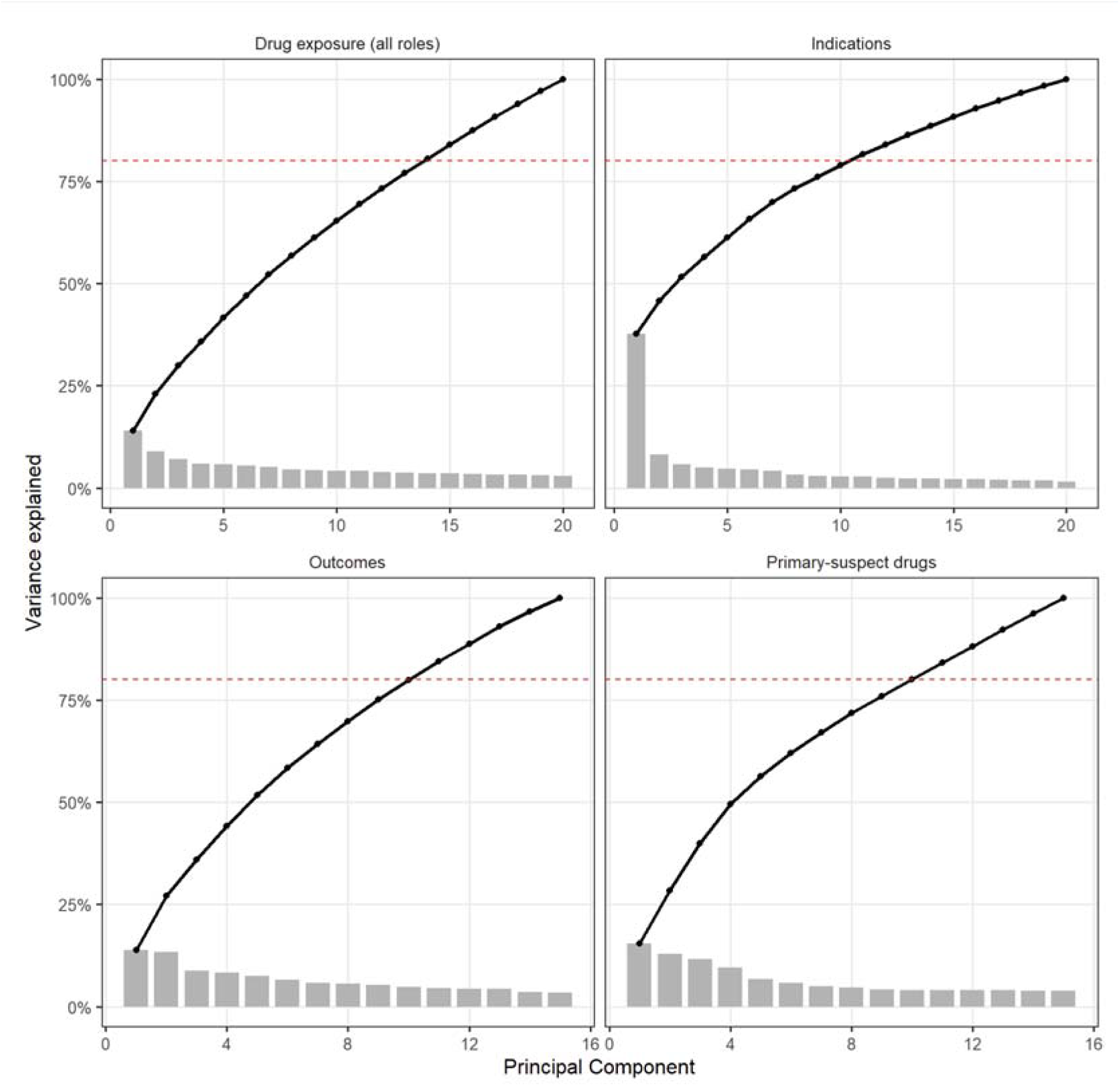
Principal component structure of drug exposures, indications, outcomes, and primary-suspect medications. Scree plots demonstrating the proportion of variance explained by principal components derived from four high-dimensional feature matrices: all reported drug exposures, indications, adverse event outcomes, and primary-suspect drugs. The dashed red line denotes 80% cumulative variance explained. Across all feature sets, variance was concentrated within a limited number of principal components, with approximately 8–14 components capturing 80% of total variance.

**Figure S4.**
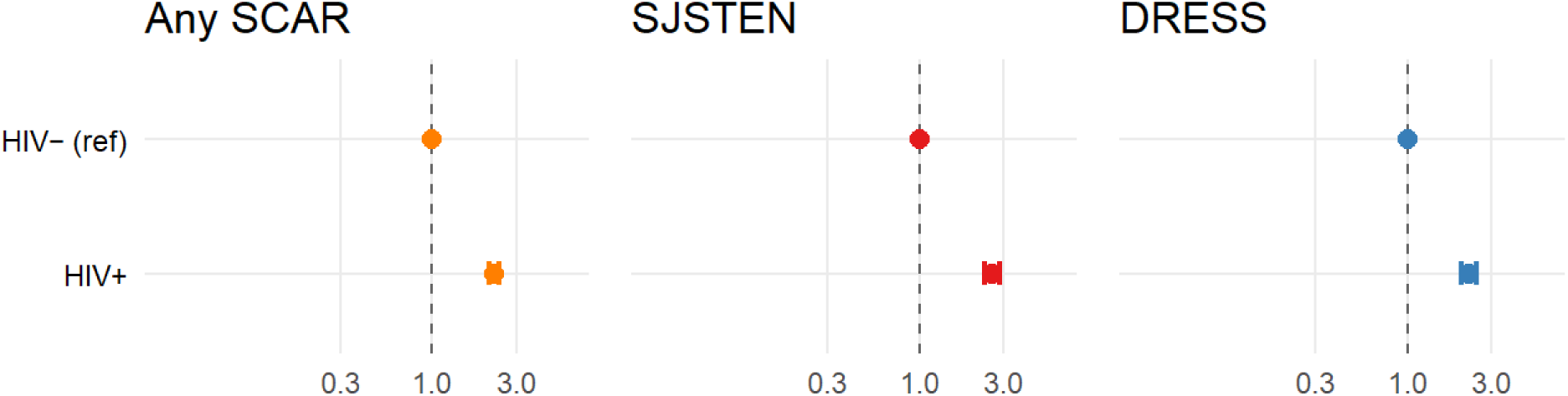
Unadjusted association between HIV infection and severe cutaneous adverse reactions. HIV infection was associated with increased odds of any SCAR (OR 2.26, 95% CI [2.11-2.41]), SJS/TEN (OR 2.57 [2.33-2.82]), and DRESS (OR 2.23 [2.46-2.82]). Estimates represent crude associations without adjustment for demographic characteristics, reporting patterns, medication burden, or drug exposure structure.

**Figure S5.**
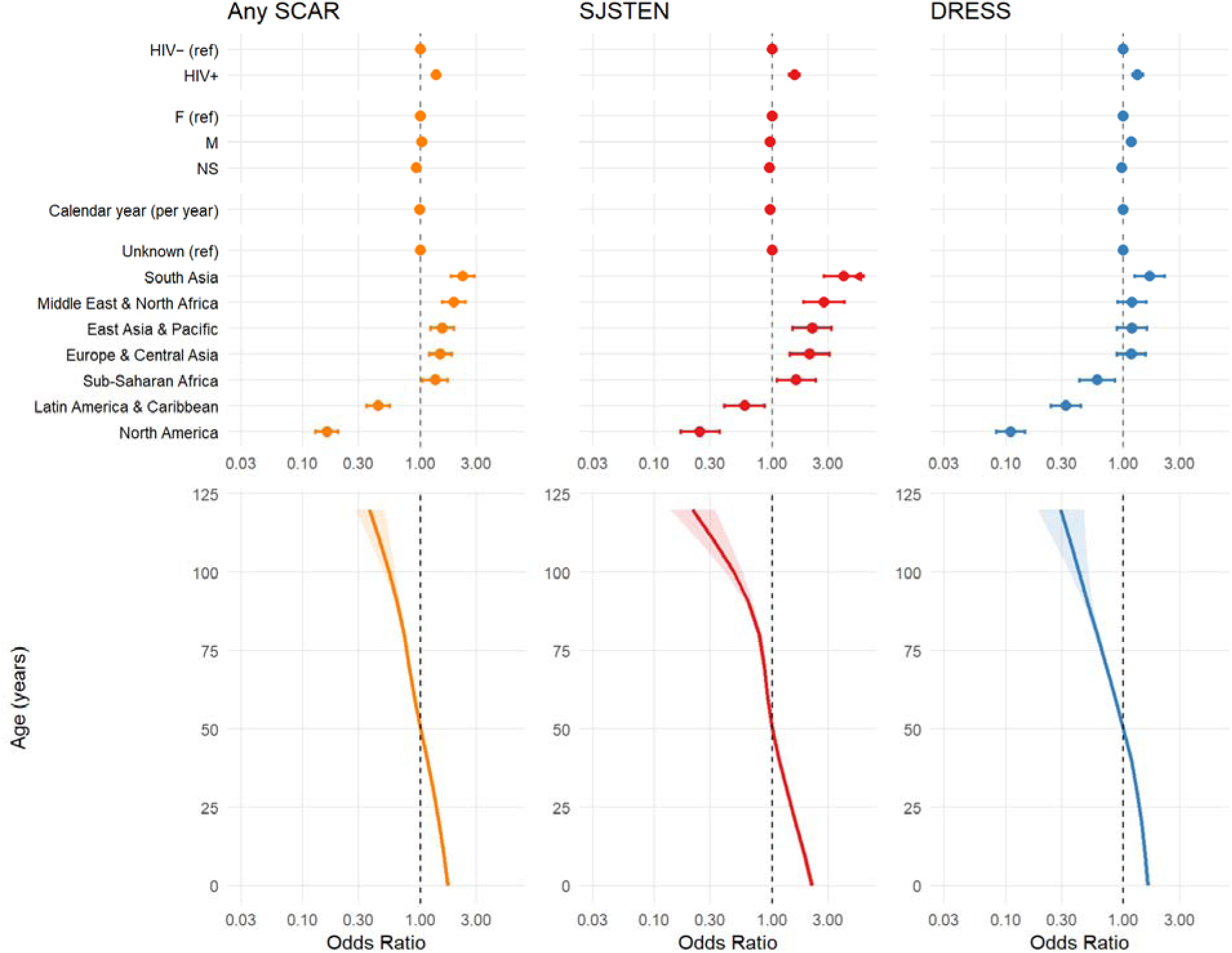
Demographic-adjusted association between HIV infection and severe cutaneous adverse reactions. Models were adjusted for age (natural cubic spline with 4 degrees of freedom), sex, geographic region, and calendar year. HIV infection remained associated with increased odds of all three SCAR outcomes – any SCAR (OR 1.37, 95% CI [1.28-1.47]), SJS/TEN (OR 1.55 [1.41-1.71]), and DRESS (OR 1.32 [1.20-1.46]) – after demographic adjustment, although effect estimates were attenuated relative to unadjusted analyses.

**Figure S6.**
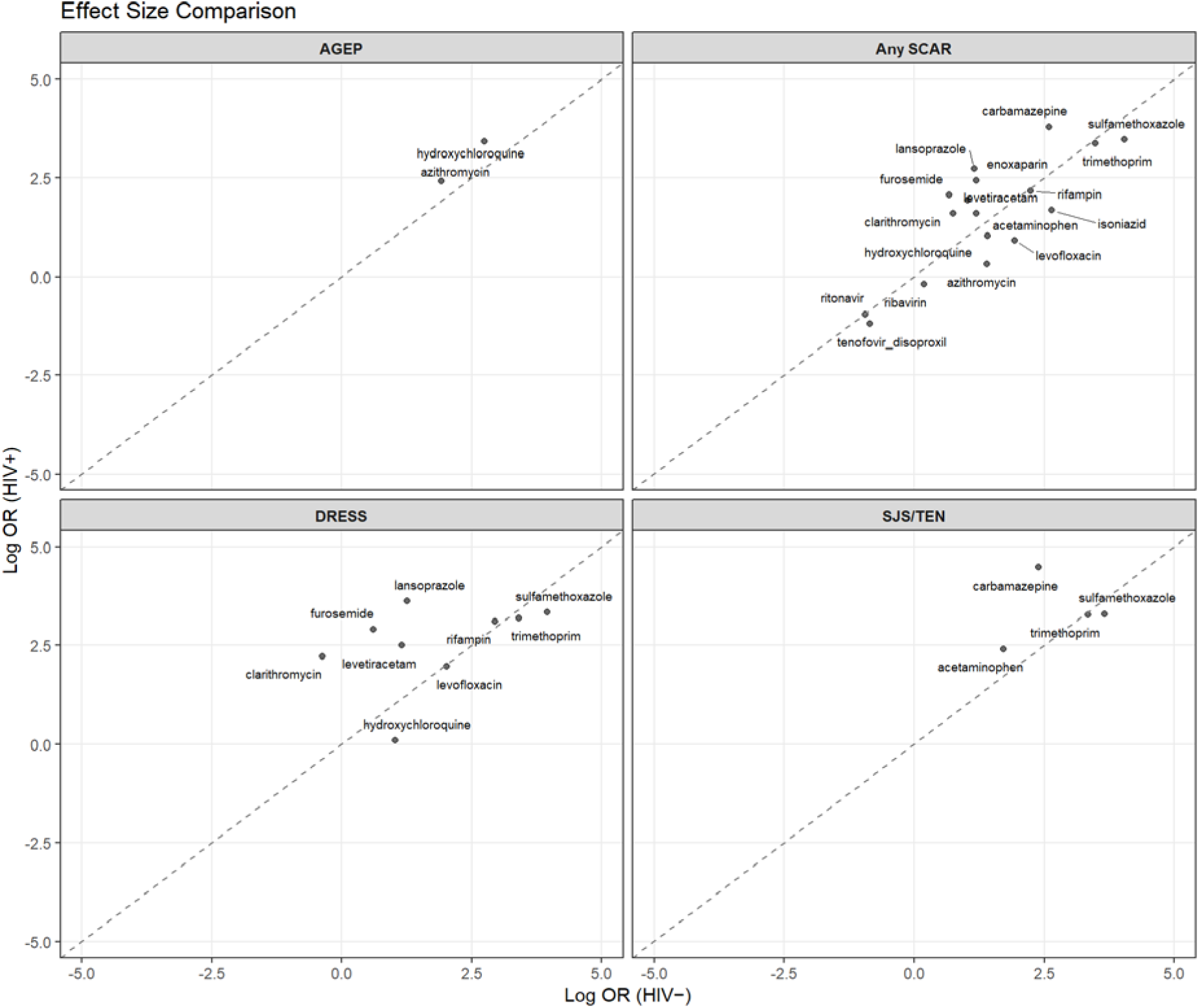
Comparison of drug-associated effect sizes in HIV-positive and HIV-negative individuals. Scatterplots comparing drug-specific RORs in HIV-negative (x-axis) and HIV-positive (y-axis) individuals for AGEP, any SCAR, DRESS, and SJS/TEN. Each point represents a primary suspect drug. The dashed diagonal line indicates equal effect sizes across HIV strata. Most drugs clustered near the line of identity, indicating similar effect sizes in HIV-positive and HIV-negative individuals, while a subset of drugs demonstrated stronger associations among HIV-positive individuals, particularly for DRESS and SJS/TEN.

**Table S7.**
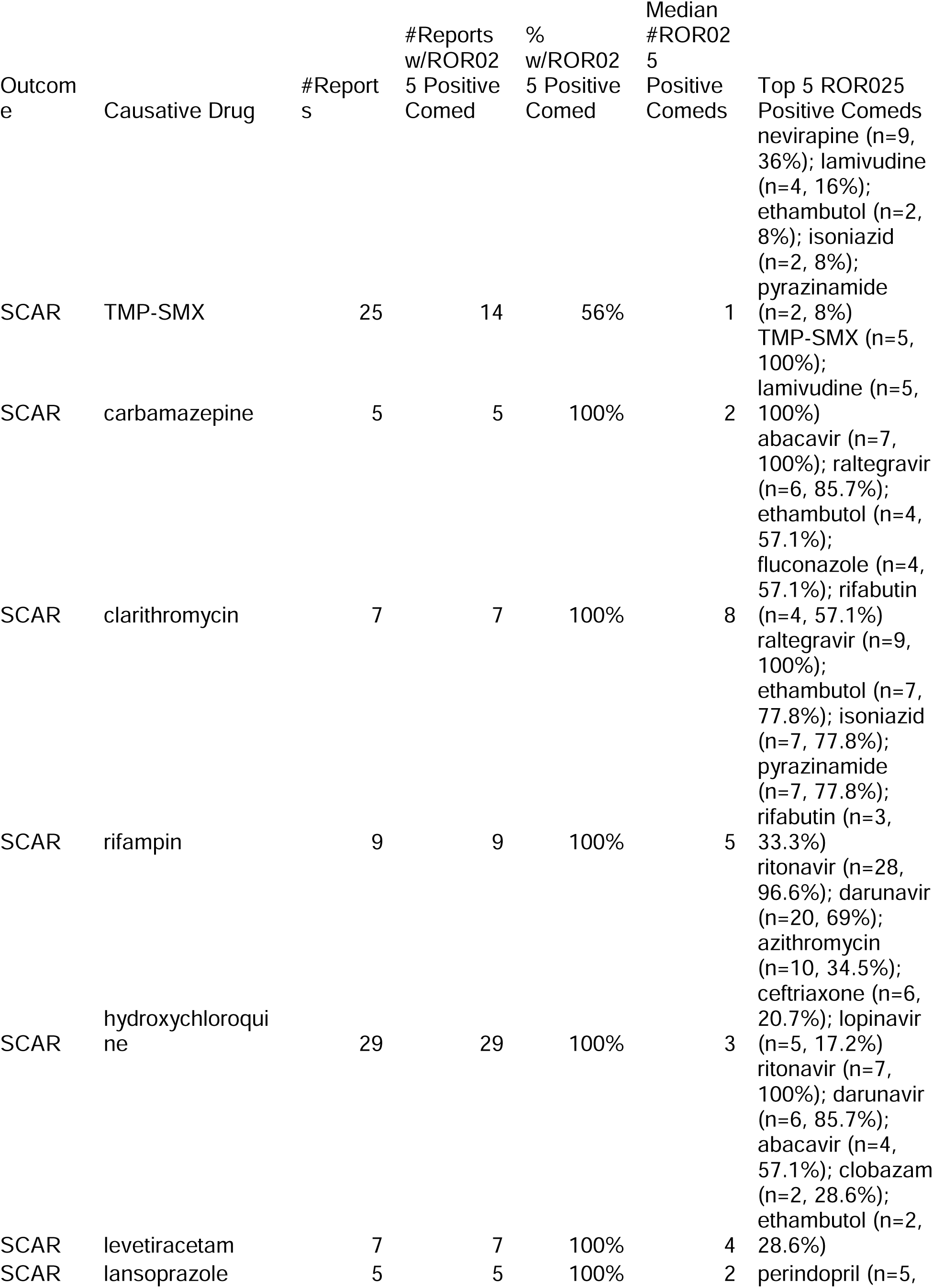
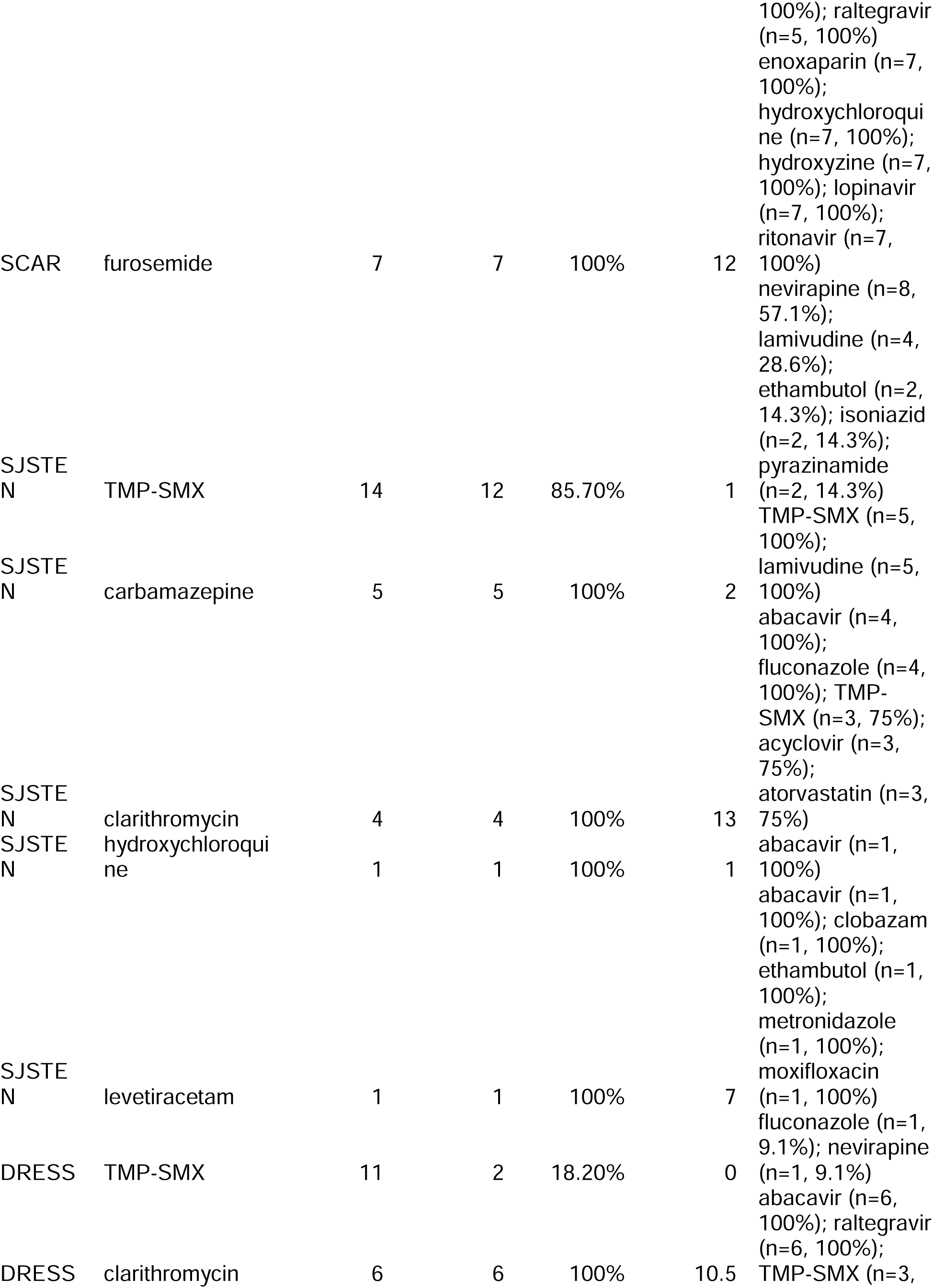
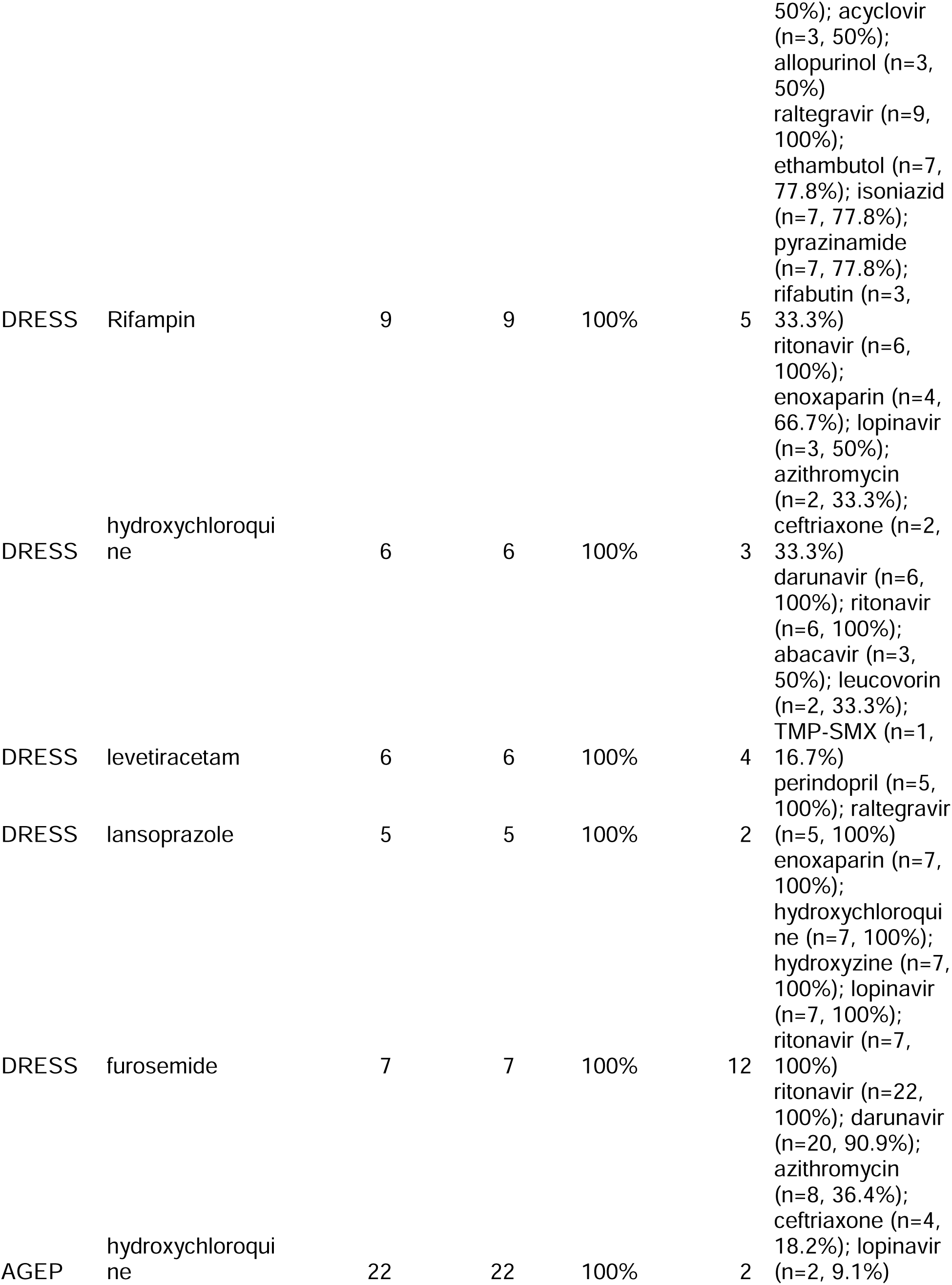
Co-medication burden among HIV-positive SCAR reports involving selected non-antiretroviral primary suspect drugs.

